# Symptom-based phenotype discovery in motor neuron disease using natural language processing of electronic health records

**DOI:** 10.64898/2026.06.18.26355960

**Authors:** Yusuf Abdulle, Vlad Dinu, Jinge Wu, Yunsoo Kim, Sanjay Budhdeo, Ahmad Al Khelfiat, Zhi Yao, Chris Tomlinson, Ammar Al-Chalabi, Honghan Wu, Richard Dobson, Alfredo Iacoangeli

## Abstract

**Background:** Motor neuron disease (MND) is a fatal neurodegenerative condition with significant clinical heterogeneity that is incompletely captured by existing phenotype classifications based on onset site. Electronic health records (EHRs) contain detailed symptom documentation in clinical narratives that may enable data-driven discovery of clinically meaningful patient subgroups.

**Methods:** We developed a natural language processing (NLP) pipeline using MedCAT to extract symptoms from clinical notes of 2,361 people with a confirmed diagnosis of MND at a tertiary neurology center. MND cohort confirmation used three complementary methods: clinic attendance records, text-based diagnosis detection, and NLP extraction with negation detection. Extracted symptoms were filtered to Unified Medical Language System semantic type T184 (Sign or Symptom) with removal of negated concepts. Patients were clustered using latent class analysis on binary symptom profiles. Survival differences were assessed using Kaplan-Meier analysis, log-rank tests, and Cox proportional hazards regression.

**Results:** From the first clinical notes, we identified four clusters of symptoms among 872 patients and 76 symptoms: Motor-Bulbar (n=373), Motor-Tremor (n=154), Sensory-Pain (n=222), and Motor-Respiratory (n=123). When extended to all clinical notes (n=2,065; 184 symptoms), these reorganized into three clusters: Autonomic-Respiratory (n=472), Nocturnal-Respiratory (n=338), and Classic Motor (n=1,255). Survival differences were significant across all clusters in both the first notes and all notes analyses (log-rank p < 0.001).

**Conclusions:** NLP-based symptom extraction from EHRs identifies clinically meaningful MND subgroups that extend beyond traditional onset-site classifications. Autonomic-respiratory symptom burden is associated with poorer survival while a newly identified Sensory-Pain subtype with a better prognosis. These data-driven phenotypes may improve prognostication and inform targeted supportive care.

## Introduction

Motor neuron disease (MND) is a fatal neurodegenerative disease characterized by progressive degeneration of upper and lower motor neurons, leading to muscle weakness, atrophy, and eventual paralysis [1]. The most common form, amyotrophic lateral sclerosis (ALS), accounts for most cases and is characterized by involvement of both upper and lower motor neurons. The disease leads to progressive weakness, muscle atrophy, and ultimately respiratory failure, with a median survival of 3–4 years from symptom onset [2]. MND carries a significant burden both for patients and healthcare systems, and despite decades of research, only two disease-modifying therapies, riluzole and edaravone, have demonstrated modest survival benefits, with no curative treatment available [1, 2]. A defining characteristic of MND is its complex and heterogenic genetic and molecular bases which, despite consistent advances, remain largely unknown [2, 3, 4, 5]. Similarly, significant clinical heterogeneity exists in disease presentation, progression rate, and survival outcomes [2].

Traditional MND phenotyping relies primarily on the site of symptom onset (bulbar vs. limb-onset) and the relative involvement of upper versus lower motor neurons [6]. Bulbar onset, which presents with dysarthria and dysphagia, is associated with shorter survival compared to limb-onset disease, while rarer phenotypes such as flail arm and primary lateral sclerosis carry distinct prognostic profiles [6]. While clinically useful, these classifications explain only a fraction of the observed variability in patient outcomes. The El Escorial criteria and their revisions provide diagnostic certainty levels but were primarily designed for clinical trial recruitment and research standardization, rather than prognostic stratification. More recently, the Gold Coast criteria have been proposed to simplify the diagnostic process while maintaining clinical utility [7]. The ENCALS prediction model and its implementation as the TRICALS risk score in clinical trials accounts for prognosis using variables including age at onset, site of onset, diagnostic delay, forced vital capacity, and progression rate [8, 9]. However, despite these criteria and models, there remains considerable unexplained prognostic variance. Further adding neurofilament light chain levels can improve prognostic accuracy, but substantial inter-individual variability remains unexplained [8, 9]. This suggests that current phenotyping approaches may not fully capture the biological and clinical complexity of MND, and that data-driven approaches to subgroup identification may reveal novel, clinically meaningful patient clusters [10, 11]

Electronic health records (EHRs) represent a rich and largely underutilized source of longitudinal clinical data. EHRs contain detailed documentation of patient symptoms, neurological examination findings, functional decline, and treatment responses accumulated over the disease course [12]. Crucially, much of this information exists in unstructured free-text clinical narratives including clinic letters, discharge summaries, and multidisciplinary team notes, that are not captured in structured coded fields and are therefore inaccessible to conventional database queries. Natural language processing (NLP) methods can extract structured clinical concepts from such unstructured text at scale, enabling large-scale phenotypic analyses that would be infeasible through manual chart review [13]. The application of NLP to EHR data has demonstrated utility across a range of neurological conditions, including cognitive decline and dementia, though its application to MND remains limited [13]. Previous work using EHRs in MND has primarily focused on structured data, and therefore the use of NLP provides a novel opportunity to uncover more information hidden in health records [14]

MedCAT (Medical Concept Annotation Toolkit) is an open-source clinical NLP tool that identifies medical entities in free text and maps them to standardized terminologies, including the Unified Medical Language System (UMLS) and SNOMED-CT [15]. UMLS semantic types allow systematic filtering of extracted concepts to specific clinical categories, such as signs and symptoms (T184), enabling targeted feature extraction for downstream analyses. Critically, MedCAT’s meta-annotation capabilities provide negation and temporality detection, distinguishing affirmed from negated findings and historical from current mentions which is a key requirement for accurate clinical phenotyping from routine clinical text.

Together, NLP-derived symptom profiles and unsupervised clustering methods offer a complementary approach to traditional phenotyping, enabling the discovery of patient subgroups defined by co-occurring clinical features rather than a priori diagnostic categories [16]. Unlike conventional approaches that rely on structured variables or expert-defined classifications, this framework allows phenotypes to emerge directly from routine clinical documentation, capturing dimensions of disease heterogeneity that may otherwise remain hidden. In MND, where substantial prognostic and clinical variability remain unexplained, such data-driven symptom phenotyping may reveal novel disease subgroups and provide insights into disease progression that are inaccessible through existing classification systems.

The aims of this study were to: (1) develop an NLP pipeline for MND cohort identification and symptom extraction from unstructured EHR data; (2) identify data-driven patient phenotypes using unsupervised symptom-based clustering; (3) evaluate survival differences between identified phenotypes; and (4) characterize the clinical profile of each phenotype.

## Methods

### Study setting and data sources

As seen in Figure 1, we carried out a retrospective cohort study using EHR data from patients attending neurology services at King’s College Hospital, a tertiary referral center for neurological conditions, including a dedicated MND clinic. The study was approved by the KERRI Committee at King’s College London.

**Figure 1:**
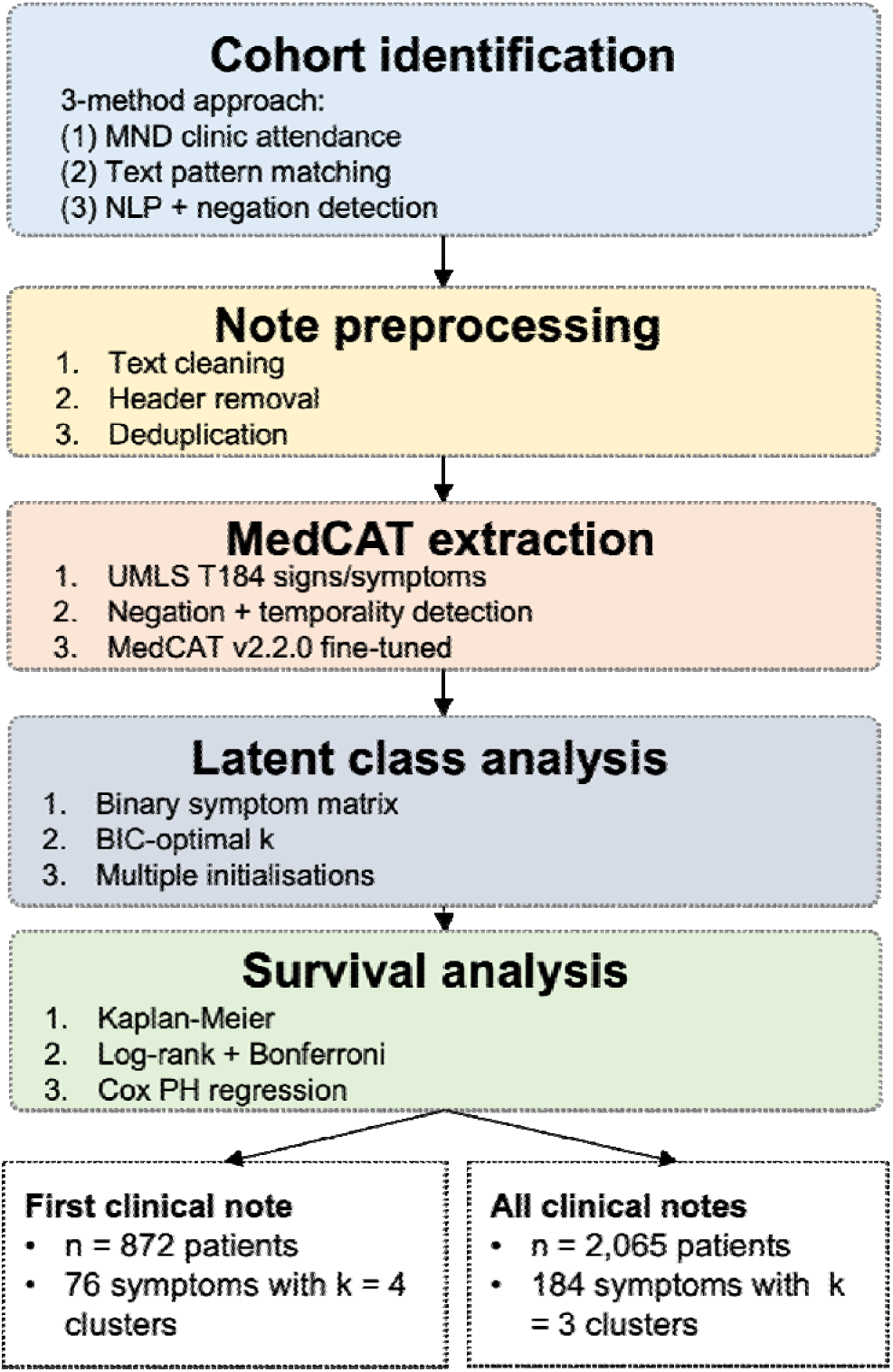
Analysis pipeline for symptom-based phenotype discovery in motor neuron disease.

Data were extracted from multiple EHR sources:

- Clinical notes and letters (outpatient clinic letters)
- Patient demographics (age, sex, dates of birth and death)
- Clinic attendance records (Patient Information Management System)

### MND cohort identification

We developed a multi-method approach combining three complementary strategies for MND cohort identification:

#### Method 1: MND clinic attendance

Patients who attended dedicated MND clinics were identified through clinic attendance records. Attendance at specialist MND services was considered strong evidence of a confirmed or highly suspected diagnosis.

#### Method 2: Text-based diagnosis detection

Clinical notes were searched for explicit diagnosis mentions using pattern matching for terms including “motor neuron disease”, “motor neuron disease”, “amyotrophic lateral sclerosis”, “ALS”, and “MND” in appropriate diagnostic context.

#### Method 3: NLP-based extraction with negation detection

We employed MedCAT to extract MND-related clinical concepts from clinical notes. MedCAT uses dictionary-based matching with contextual disambiguation to identify medical concepts and map them to UMLS Concept Unique Identifiers. We used MedCAT’s meta-annotation capabilities for negation detection, distinguishing affirmed diagnoses (“patient has MND”) from negated mentions (“no evidence of MND”) and hypothetical discussions (“query MND”). The final cohort of people confirmed diagnosis of MND included patients identified by any of the three methods.

### Clinical note preprocessing

Raw clinical notes underwent systematic preprocessing:

1. **Text cleaning:** Removal of extraneous characters, standardization of whitespace, and handling of encoding issues
2. **Header/footer removal:** Automated removal of standard letter templates, addressee information, and signature blocks using pattern matching
3. **Document length filtering:** Exclusion of extremely short (*<* 50 characters) or extremely long documents (beyond 95th percentile)
4. **Deduplication:** Removal of duplicate notes based on content similarity

### Symptom extraction using MedCAT

#### Model configuration

We used MedCAT version 2.2.0 with a UMLS-based vocabulary. The base model was fine-tuned on our clinical corpus using unsupervised training to adapt to institution-specific terminology, abbreviations, and documentation patterns.

#### Concept extraction and filtering

Medical concepts were extracted from all clinical notes for patients confirmed diagnosis of MND. To focus on symptoms relevant to phenotyping, we applied the following filters:

- **Semantic type filter:** Retained only concepts with UMLS semantic type T184 (“Sign or Symptom”)
- **Negation filter:** Removed concepts identified as negated by MedCAT’s meta-annotation model
- **Confidence filter:** Excluded low-confidence extractions (context similarity *<* 0.5)
- **Specificity filter:** Removed overly generic terms (e.g., “symptom”, “finding”, “problem”)

#### Patient-symptom matrix construction

For each patient, we created a binary feature vector indicating the presence (1) or absence (0) of each symptom across all their clinical notes. Symptoms documented in fewer than 5 patients were excluded to reduce noise from rare or potentially erroneous extractions. The resulting patient-symptom matrix had dimensions *n* × *m*, where *n* is the number of patients and *m* is the number of retained symptoms.

### Clustering methodology

#### Algorithm selection

Patients were clustered based on symptom profiles using latent class analysis (LCA), a probabilistic model-based approach suited to binary feature matrices [17]. LCA models each latent class as a product of independent Bernoulli distributions, estimating per-class symptom probabilities via expectation-maximization. Multiple random initializations (*n* = 10) were used to avoid local optima, retaining the solution with the highest log-likelihood.

#### Optimal cluster selection

The optimal number of clusters was determined by evaluating *k* ∈ {2, 3*, …,* 10} using:

- **Bayesian Information Criterion (BIC):** Balancing model fit against complexity, with lower BIC indicating better model selection
- **Akaike Information Criterion (AIC):** As a complementary criterion for model comparison

The final cluster number was selected based on BIC minimization with consideration of clinical interpretability.

#### Visualization

Dimensionality reduction using UMAP with Jaccard distance was performed for exploratory visualization (Supplementary Information), though we note that UMAP projections are not expected to separate clusters identified by latent class analysis, which captures probabilistic symptom co-occurrence patterns rather than geometric distance in feature space.

#### Cluster characterization

Each cluster was characterized by identifying symptoms with the highest within-cluster prevalence relative to overall cohort prevalence. The top 10 discriminating symptoms per cluster were reported.

### Survival analysis

#### Outcome definition

The primary outcome was overall survival, defined as time from first recorded clinical encounter (proxy for diagnosis date) to death from any cause. Patients alive at study end or lost to follow-up were censored at their last recorded encounter.

#### Patient and Public Involvement

This study was conducted using retrospective, routinely collected EHR data and the research questions addressed such as improving phenotypic characterization, prognostic stratification, and care planning in MND was identified in close dialogue with the clinical MND multidisciplinary team at King’s College Hospital, as well as patient groups, whose expertise reflects the priorities and unmet needs of patients living with MND. Dissemination of findings to patient communities is planned as part of subsequent work. This work was conducted in accordance with the Declaration of Helsinki.

#### Statistical methods

**Kaplan-Meier analysis:** Survival curves were estimated for each cluster. Median survival with 95% confidence intervals was calculated.

**Log-rank tests:** Overall comparison between clusters used the multivariate log-rank test. Pairwise comparisons employed Bonferroni correction.

**Cox proportional hazards regression:** Cox models estimated hazard ratios (HRs) with 95% CIs for time from first clinical encounter to death or censoring (event = all-cause mortality). Models were adjusted for age at first encounter, sex (male vs. female), and ethnicity (categorical). Cluster assignment was entered as a single categorical predictor (*k* levels; reference = lowest-mortality cluster); an overall likelihood-ratio test assessed whether cluster affected survival, with cluster-specific HRs reported versus the reference. Within each cluster, separate models tested binary symptom presence (MedCAT-derived), requiring ≥10 affected patients; HRs were not estimable when symptoms were near-universal within a cluster. The proportional hazards assumption was assessed using Schoenfeld residuals. Additional analyses, including individual symptom hazard ratios, symptom combination analysis, and diagnostic delay sensitivity analysis, are described in the Supplementary Information.

## Results

### Cohort characteristics

The cohort of patients with a confirmed diagnosis of MND comprised 2,361 patients. Table 1 summarizes baseline characteristics.

**Table 1:** Baseline characteristics of the MND cohort.

| Characteristic | Value |
| --- | --- |
| Total patients, n | 2,361 |
| Deaths, n (%) | 1,520 (64.4%) |
| **Demographics** |  |
| Age, mean (SD) years | 60.8 (13.4) |
| Median age (IQR) years | 62.0 (54.0–71.0) |
| Male, n (%) | 1,435 (60.8%) |
| Female, n (%) | 919 (38.9%) |
| **Ethnicity** |  |
| White, n (%) | 1,207 (51.1%) |
| Unknown, n (%) | 1,008 (42.7%) |
| Black, n (%) | 96 (4.1%) |
| Asian, n (%) | 21 (0.9%) |

### Symptom extraction

MedCAT extracted symptoms from clinical notes for all patients with a confirmed diagnosis of MND. From the first clinical notes, 76 unique symptoms were identified among 872 patients (Supplementary Table 2 reports the complete list of symptoms). This minimum symptom threshold was applied to both analyses to ensure that cluster assignments were based on sufficiently informative symptom profiles rather than sparse or near-empty records, which would introduce noise into the latent class model. In the first-note analysis, most excluded patients had initial clinic letters that were brief administrative summaries or referral acknowledgements containing fewer than five extractable symptom concepts, rather than substantive clinical assessments. The all-notes analysis retains a substantially larger proportion of the cohort (2,065 of 2,361; 87.5%) because the cumulative record across multiple encounters almost always yields sufficient symptom documentation to meet the inclusion threshold. The two analyses should therefore be understood as complementary rather than directly comparable: the first-note analysis characterizes early presentation phenotypes in patients with informative initial documentation, while the all-notes analysis represents the broader MND population seen at this center.

From all available clinical documentation, 184 unique symptoms were extracted from 2,065 patients, after filtering to UMLS semantic type T184 (Sign or Symptom) and removing negated concepts (Supplementary Table 3 reports the complete list of symptoms). It is important to note that synonyms here are merged through MedCAT. For example, if a patient were to report twitching, the MedCAT tool would tag this as fasciculations. Figure 2 shows the distribution of the top 20 most common symptoms for all patients in our dataset.

**Figure 2:**
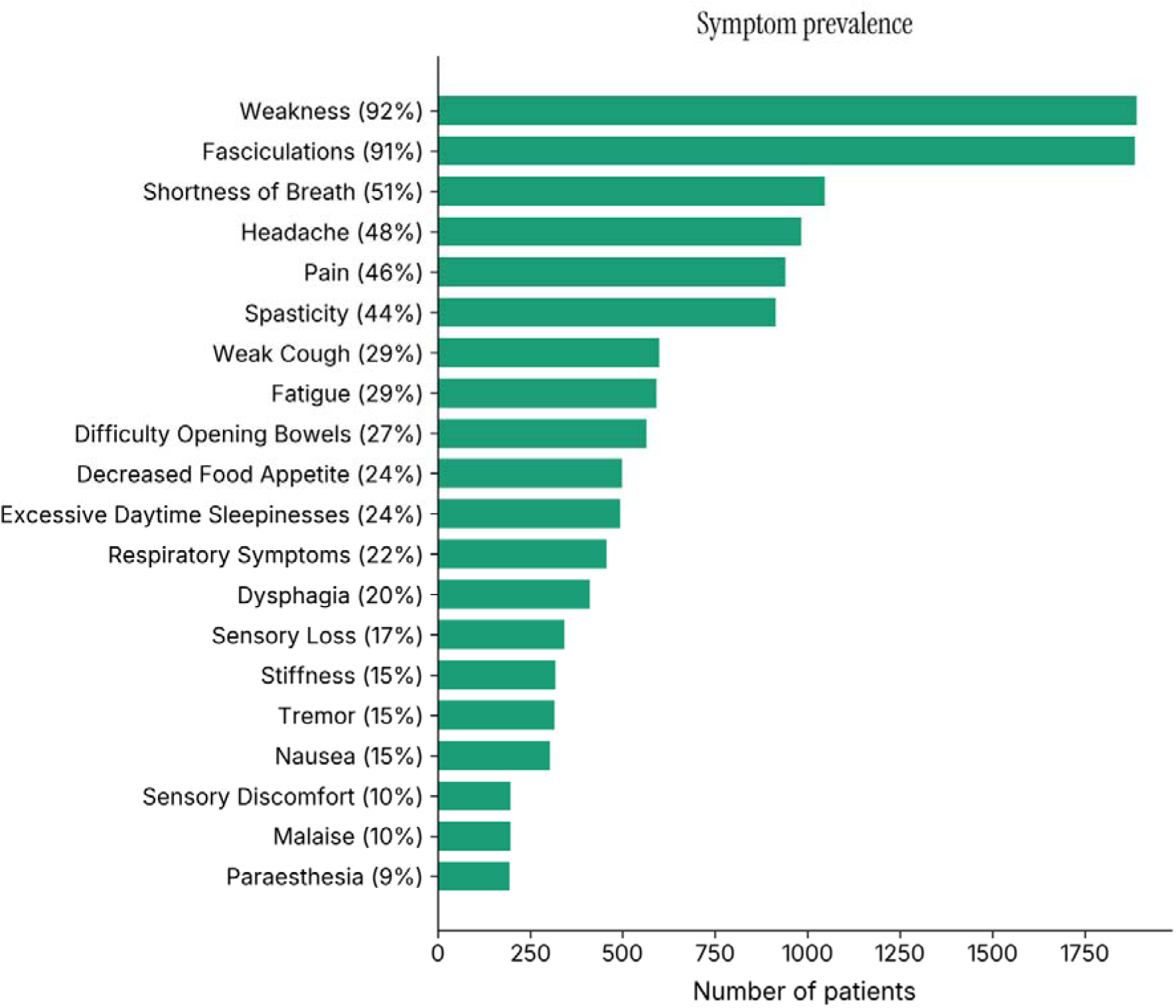
Most frequently extracted symptoms across the MND cohort. Bars represent number of patients with each symptom documented. General weakness and fasciculations were near-universal. Symptoms such as tremor and sensory loss appear; symptoms typically not regarded as MND symptoms.

### Initial phenotypes: analysis of first clinical notes

We first analyzed symptoms documented in the initial clinical note for each patient. Latent class analysis with BIC-based model selection identified four optimal clusters (*n* = 872; 76 symptoms). BIC was evaluated for *k* = 2 to *k* = 10, with the minimum at *k* = 4.

#### First-note cluster characteristics

The four clusters identified showed distinct clinical phenotypes. Most frequent symptoms and clinical characteristics of the four clusters are reported in Table 2.

**Table 2:** Symptom-based cluster characteristics and survival outcomes from first clinical notes (*n* = 872)

| Cluster Phenotype | *n* | % | Male, *n* (%) / Female, *n* (%) | Mean age at diagnosis, (yrs) (SD) | Median Survival (years) (IQR) | Defining symptoms |
| --- | --- | --- | --- | --- | --- | --- |
| Motor-Bulbar | 373 | 42.8 | 243 (65.1) / 129 (34.6) | 61.1 (±13.2) | 1.80 (0.93–4.55) | Weakness (91%), fasciculations (89%), Spasticity (28%)<br>dyspnoea (27%), dysphagia (20%) |
| Motor-Tremor | 154 | 17.7 | 99(64.3) / 55 (35.7) | 61.3 (±13.8) | 1.98 (0.82–4.87) | Fasciculations (100%), weakness (75%), tremor (47%), spasticity (34%), slow movement (16%) |
| Sensory-Pain | 222 | 25.5 | 143 (64.4) / 78 (35.1) | 58.6 (±14.5) | 3.82 (1.38–9.25) | Fasciculations (82%), weakness (82%), pain (52%), sensory loss (41%), paraesthesia (16%) |
| Motor-Respiratory | 123 | 14.1 | 72 (58.5) / 50 (40.7) | 61.2 (±15.1) | 2.51 (1.06–3.75) | Dyspnoea (100%), excessive daytime sleepiness (99%), decreased appetite (97%), weak cough (94%), Spasticity (94.3%) |
| **Total** | **872** | **100** |  |  |  |  |

#### First-note survival analysis

Kaplan-Meier analysis showed significant survival differences between the four first-note clusters (log-rank *p <* 0.001; Figure 3)

**Figure 3:**
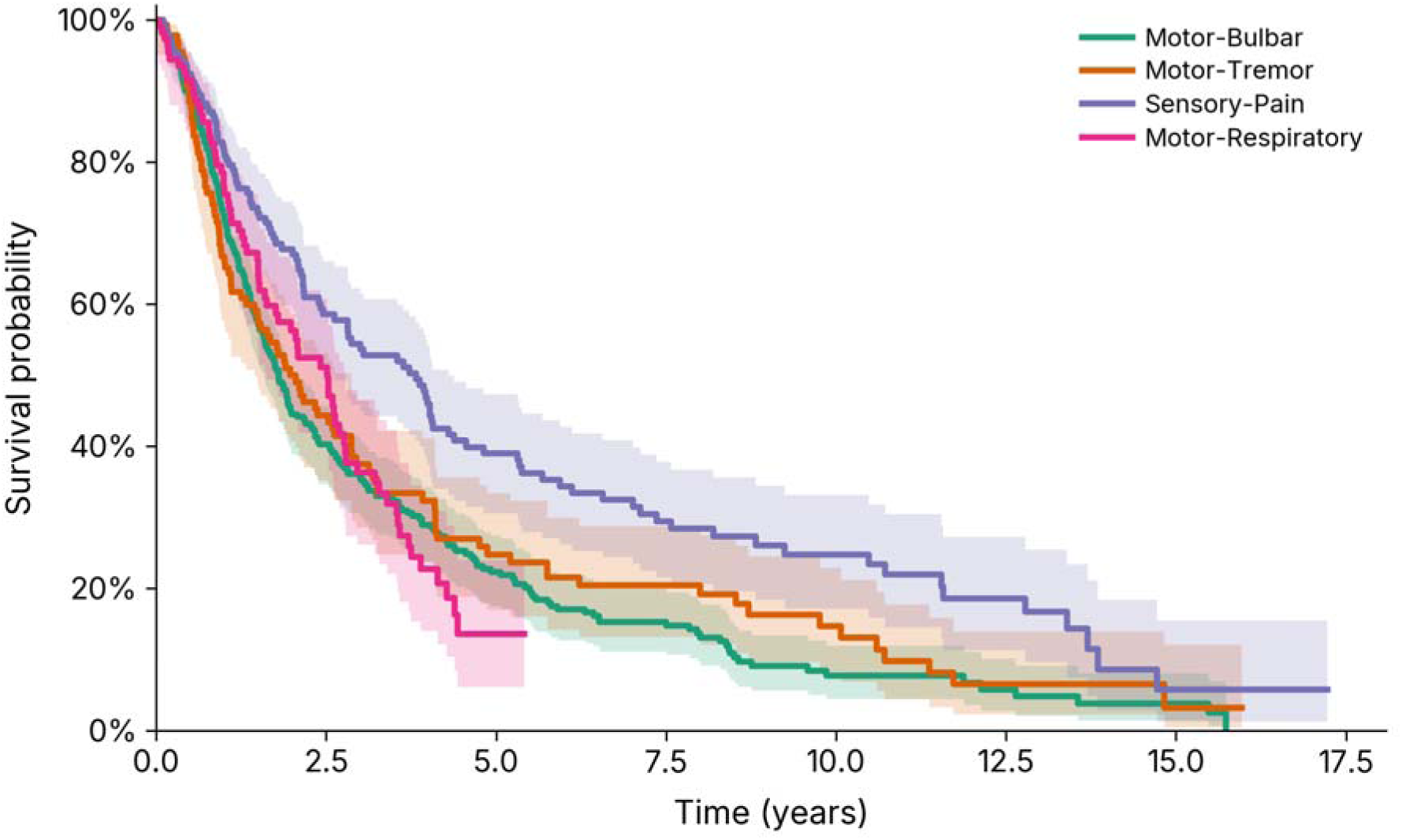
Kaplan-Meier survival curves by cluster for first clinical note analysis (*n* = 872). Curves show cumulative survival over time for each of the four identified phenotypes. Log-rank test: *p <* 0.001.

However, not all clusters showed significant survival differences with the others when compared individually to all other clusters using Bonferroni correction (α = 0.05*/*6 = 0.0083, Table 3).

**Table 3:** Pairwise log-rank tests, first clinical note clusters.

| Clusters | Phenotype Comparison | $X^2$ | Corrected p-value | Sig. |
| --- | --- | --- | --- | --- |
| 0 vs. 2 | Motor-Bulbar vs. Sensory-Pain | 19.11 | <0.05 | Yes |
| 1 vs. 2 | Motor-Tremor vs. Sensory-Pain | 8.58 | <0.05 | Yes |
| 2 vs. 3 | Sensory-Pain vs. Motor-Respiratory | 11.63 | <0.05 | Yes |
| 0 vs. 1 | Motor-Bulbar vs. Motor-Tremor | 0.53 | >0.05 | No |
| 0 vs. 3 | Motor-Bulbar vs. Motor-Respiratory | 0.004 | >0.05 | No |
| 1 vs. 3 | Motor-Tremor vs. Motor-Respiratory | 0.135 | >0.05 | No |

The Motor-Bulbar, Motor-Tremor, and Motor-Respiratory clusters did not have statistically different survival (*p >* 0.05), while Sensory-Pain had a significantly better prognosis than each of the other clusters. Cox analysis aligns with the results of the Kaplan-Meier analysis (Supplementary Table 4). Supplementary Tables 6–9 report the association between survival and the five most frequent symptoms within each phenotype cluster.

### Extended phenotypes: analysis of all clinical notes

We repeated the clustering analysis using all documented symptoms across each patient’s entire clinical record (2,065 patients, 184 symptoms). Latent class analysis with BIC-based model selection identified *k* = 3 as optimal. The reduction from four to three clusters indicates that initial presentation heterogeneity reorganized into three core phenotypic patterns when the complete clinical history is considered, seen in Table 4 (see Supplementary Table 3 for more extensive list of characterizing symptoms).

**Table 4:** Symptom-based cluster characteristics and survival outcomes from all clinical notes (*n* = 2,065)

| Cluster Phenotype | *n* | % | Male, *n* (%) / Female, *n* (%) | Mean age at diagnosis (yrs) (SD) | Median survival (years) (IQR) | Defining symptoms |
| --- | --- | --- | --- | --- | --- | --- |
| Autonomic-Respiratory | 472 | 22.9 | 303 (64.2) / 169 (35.8) | 60.2 (±12.4) | 3.15 (1.54–7.28) | Weakness (98%), fasciculations (95%), headache (82%), pain (73%), dyspnoea (85%) |
| Nocturnal-Respiratory | 338 | 16.4 | 203 (60.1) / 133 (39.3) | 61.2 (±14.3) | 2.51 (1.13–6.51) | Dyspnoea (100%), daytime sleepiness (100%), morning headache (98%), decreased appetite (98%), weak cough (96%) |
| Classic Motor | 1,255 | 60.8 | 767 (61.1) / 484 (38.6) | 61.7 (±13.6) | 1.94 (0.93–4.51) | Fasciculations (91%), Weakness (90%), pain (34%), spasticity (30%), shortness of breath (25.3%) |
| **Total** | **2,065** | **100** |  |  |  |  |

#### All-notes survival analysis

Kaplan-Meier analysis showed significant survival differences between the three clusters (*p <* 0.001; Figure 4).

**Figure 4:**
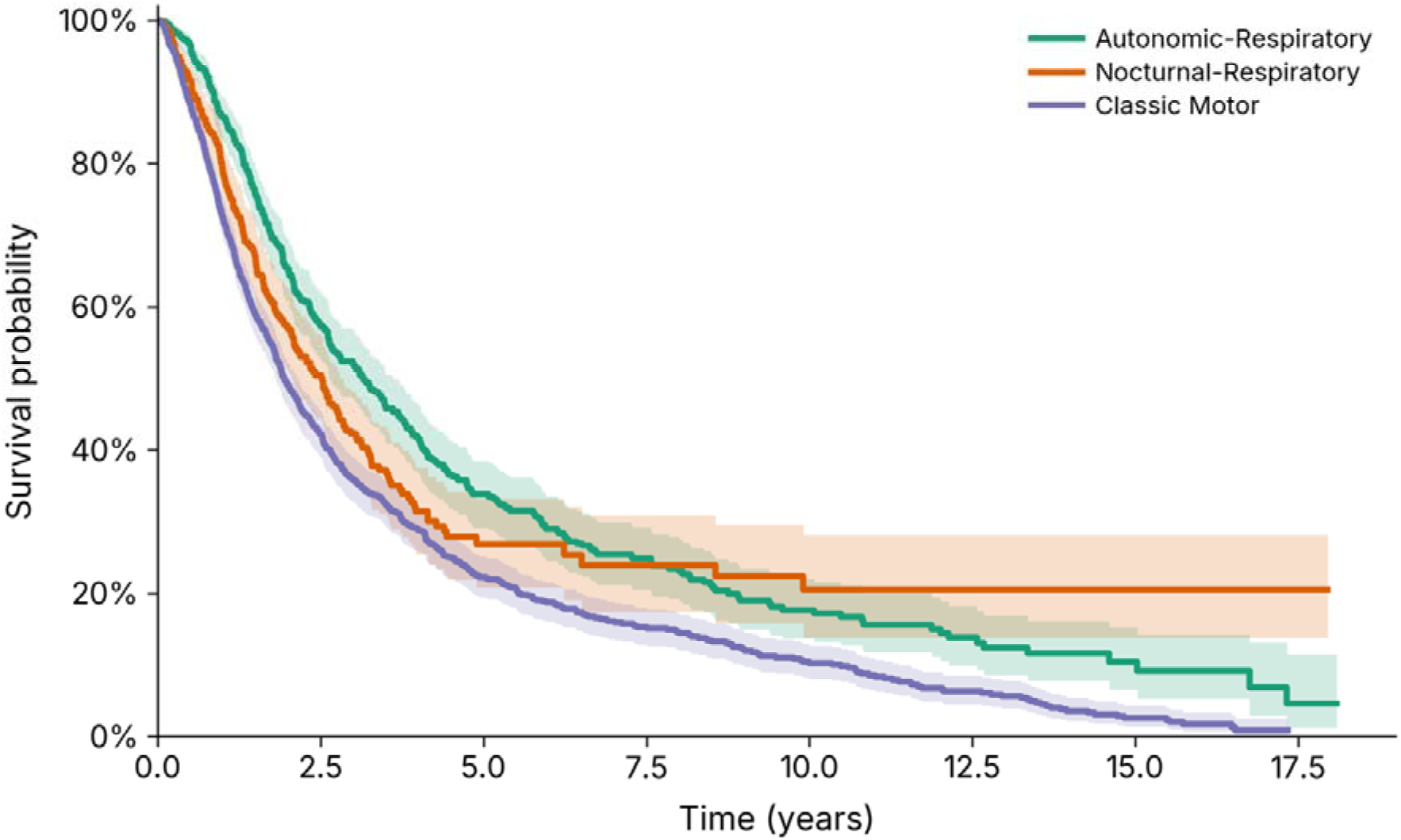
Kaplan-Meier survival curves by cluster for all clinical notes analysis (*n* = 2, 065). Curves show cumulative survival over time for each of the three identified phenotypes. Log-rank test: *p <* 0.001.

Pairwise log-rank comparisons (Table 5) with Bonferroni correction (α= 0.05*/*3 = 0.0167): Autonomic-Respiratory and Classic Motor clusters showed significant survival differences (*p <* 0.001), as did Nocturnal-Respiratory versus Classic Motor (*p* = 0.001). The Nocturnal-Respiratory cluster did not differ significantly from Autonomic-Respiratory (*p* = 0.078). Cox analysis aligns with the results of the Kaplan-Meier analysis (supplementary table 5). Supplementary Tables 6–9 report the association between survival and the five most frequent symptoms within each phenotype cluster.

**Table 5:** Pairwise log-rank tests, all clinical notes clusters.

| Clusters | Phenotype comparison | $\chi^2$ | p-value | Sig. |
| --- | --- | --- | --- | --- |
| 0 vs. 2 | Autonomic-Respiratory vs. Classic Motor | 39.54 | <0.05 | Yes |
| 1 vs. 2 | Nocturnal-Respiratory vs. Classic Motor | 10.79 | <0.05 | Yes |
| 0 vs. 1 | Autonomic-Respiratory vs. Nocturnal-Respiratory | 3.10 | >0.05 | No |

#### Survival differences between clusters

Median survival varied across clusters in both analyses (Figure 5). In the first-note analysis, the Sensory-Pain cluster had the longest median survival (3.82 years), compared to Motor-Respiratory (2.51 years) Motor-Tremor (1.98 years), and Motor-Bulbar (1.80 years). The three motor-predominant clusters did not differ significantly from one another (all pairwise log-rank p > 0.05 after Bonferroni correction), while the Sensory-Pain cluster had significantly longer survival than each of the remaining three (all p < 0.0083). In the all-notes analysis, the Autonomic-Respiratory cluster had the longest median survival (3.15 years), followed by Nocturnal-Respiratory (2.51 years) and Classic Motor (1.94 years). Autonomic-Respiratory and Classic Motor clusters differed significantly (p < 0.001), as did Nocturnal-Respiratory and Classic Motor (p = 0.001), while the two respiratory clusters did not differ from one another (p = 0.078).

**Figure 5:**
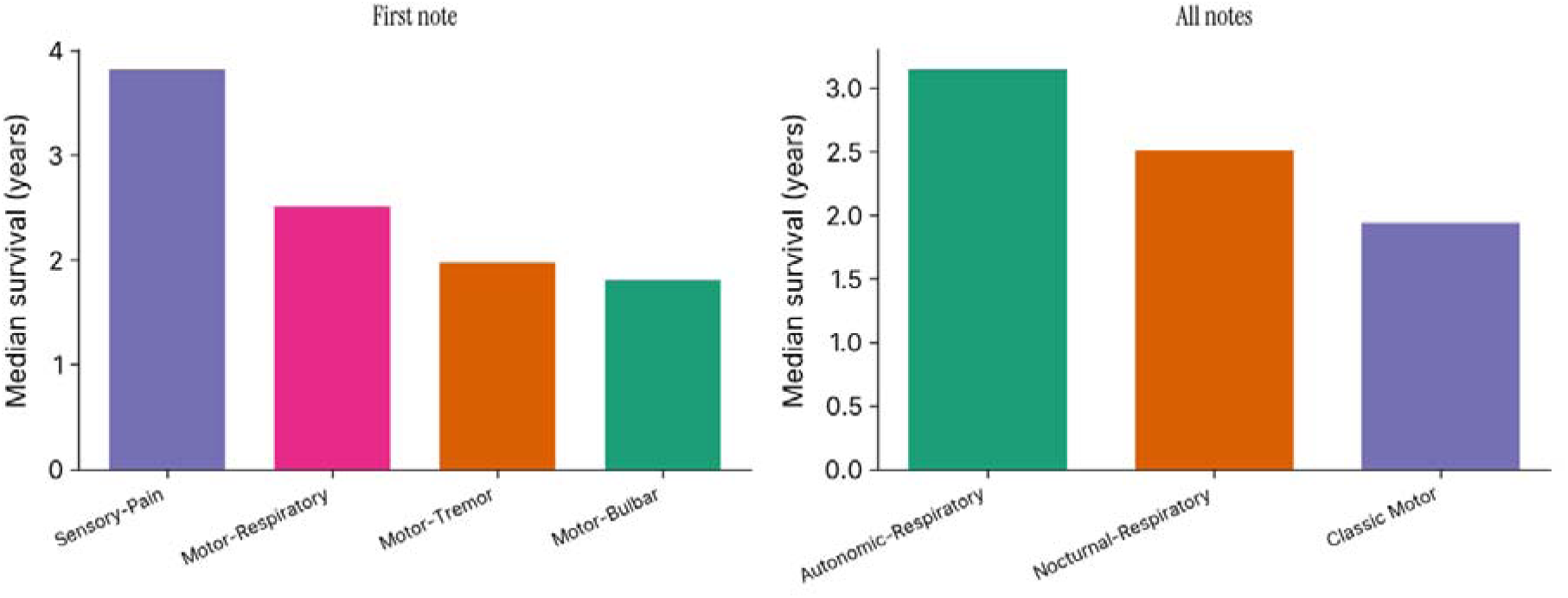
Median survival by symptom-based phenotype cluster.

### Patient flow

One phenotype, the Motor-Bulbar cluster, remains relatively stable across both analyses, with most patients retaining a similar classification. In contrast, patients assigned to the Motor-Tremor, Sensory-Pain, and Motor-Respiratory clusters at first presentation become redistributed across two broader phenotypic groups in the all-notes analysis, shown in Figure 6. This redistribution suggests that distinctions apparent at initial presentation become less pronounced when symptoms accumulated throughout the disease course are considered.

**Figure 6:**
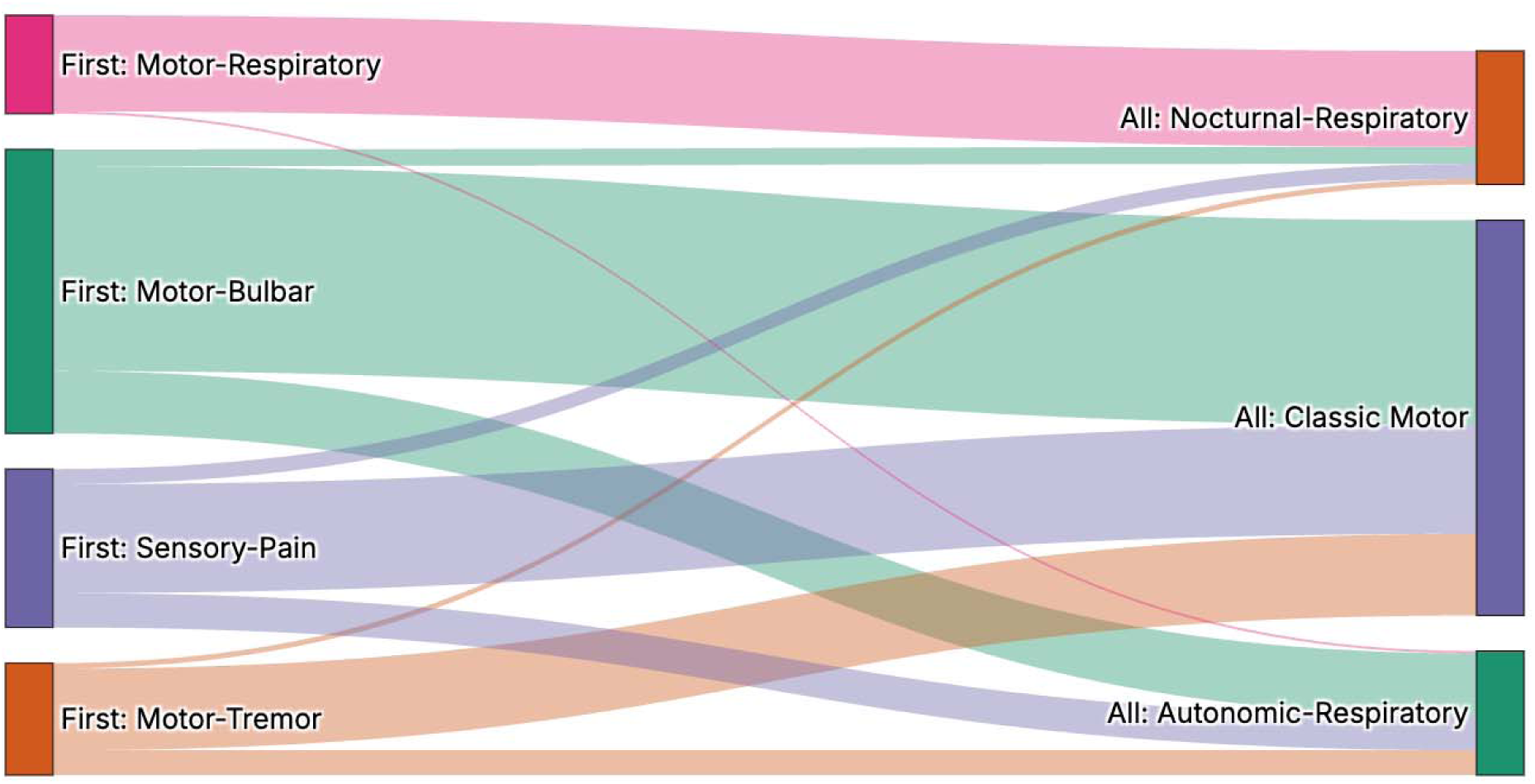
This figure illustrates how patients classified using symptoms from their first clinical note are reorganized when symptom information from the entire clinical record is incorporated. The diagram includes only patients represented in both analyses, allowing direct comparison of phenotype assignment before and after the addition of longitudinal symptom information. When demonstrating the patient flow, we restrict the data to include only those who have viable first notes, and their progression into their respective clusters.

Overall, the figure indicates a reduction in phenotypic complexity from four initial symptom profiles to three broader patterns. Rather than revealing entirely new phenotypes, the addition of follow-up symptom information appears to increase overlap between patient groups, leading to broader and less distinct classifications. This observation is consistent with the possibility that patients present with a more diverse spectrum of symptoms early in the disease course, while later symptom accumulation results in increasingly similar clinical profiles.

## Discussion

We developed an NLP pipeline using MedCAT to extract symptoms from clinical free-text notes and applied latent class analysis to discover data-driven phenotypes in a real-world MND cohort. Analysis of first clinical notes identified four phenotypes, which reorganizes into three broader phenotypes when extended to all available clinical documentation across the disease course. Survival differed significantly across clusters in both analyses, suggesting that co-occurring symptom patterns carry prognostic information beyond that captured by conventional phenotyping approaches.

In the first-note approach, the UMAP projections shown in Supplementary Figures 1 showed substantial spatial overlap between the first three clusters, despite their markedly different survival trajectories. This is expected as the groups share the most common MND symptoms (weakness, fasciculations) and differ primarily in a small number of specific symptom combinations, such as bulbar involvement, respiratory compromise, and sensory-pain patterns, which make up only a small fraction of the total symptom profile. Because UMAP arranges patients by overall similarity in high-dimensional space, it cannot separate patients who look broadly alike but differ in these critical co-occurring symptoms. The fact that visually overlapping groups still show divergent survival outcomes illustrates why a model-based approach is necessary: unlike distance-based methods, latent class analysis does not group by geometric proximity but instead estimates class-specific probability distributions over symptom combinations, allowing it to detect structured co-occurrence patterns that contribute little to overall distance but carry prognostic weight [23].

### Interpretation of identified phenotypes

Looking at the most frequent symptoms occurring in the notes, tremor, a symptom that traditionally is not regarded as part of MND, is seen. It is a known phenomenon, and points towards a pathophysiological basis in MND [18]. Its presence as a cluster-defining symptom can raise the question of whether this group may include patients with overlapping or alternative diagnoses, which can mimic MND clinically and is more frequently associated with tremor. Our multi-method cohort confirmation strategy, combining MND clinic attendance records, text-based diagnosis detection, and NLP extraction with negation detection, was designed to minimize the inclusion of patients without a confirmed MND diagnosis. Nonetheless, we cannot entirely exclude the possibility that a minority of Motor-Tremor cluster patients carry a dual or alternative diagnosis. In addition, our list of the most common symptoms reveals that pain is commonly reported in the notes of patients with MND, a phenomenon that is increasingly recognized in several studies [19, 20, 21]. Finally, it is important to note that sleep deprivation has been reported to increase pain sensitivity and should be noted [22]

The observation that Classic Motor patients had the shortest median survival in the all-notes analysis, despite appearing to represent a more “typical” disease phenotype, may initially seem counterintuitive. This may reflect a survivor enrichment effect: patients in the Autonomic-Respiratory and Nocturnal-Respiratory clusters have, by definition, accumulated respiratory and autonomic symptom burden across their longitudinal record, a pattern that presupposes a sufficient period of clinical follow-up for these symptoms to emerge and be documented. Patients with very rapidly progressive disease, who may have been expected to accrue respiratory features, may die before these symptoms are extensively documented, leading to their classification within the Classic Motor group based on early-predominant motor symptoms. These considerations underscore the importance of time-aware modelling in future analyses.

The phenotypes identified in both analyses show broad alignment with established clinical subtypes of MND, providing face validity for the NLP-driven clustering approach. The Motor-Bulbar cluster corresponds to the bulbar-onset phenotype which is consistently associated with shorter survival [24]. The Motor-Respiratory cluster likely captures patients with early diaphragmatic involvement and respiratory insufficiency, a recognized poor-prognosis subgroup in whom early non-invasive ventilation assessment is critical [25]. The Motor-Tremor requires further analyses. Sensory involvement in MND has historically been considered atypical, though emerging neuropathological and electrophysiological evidence suggests that non-motor features are more common than traditionally appreciated [26, 27].

### Robustness of phenotypes: first-note to all-notes consistency

Our study also showed that the four symptom-based phenotypes identified from the first clinical notes reorganized into three broader phenotypes when symptoms from the entire clinical record were considered. One possible interpretation of this finding is that phenotypic heterogeneity is greatest early in the disease course and becomes progressively less distinct as the disease advances. Patients may initially present with diverse combinations of symptoms reflecting different patterns of anatomical involvement, rates of progression, or underlying biological mechanisms. However, as neurodegeneration progresses and symptoms accumulate, increasing overlap in clinical manifestations may lead to convergence towards a smaller number of broader phenotypic patterns.

Importantly, this observation does not necessarily imply that distinct disease subtypes disappear over time. Rather, it may reflect the fact that different disease trajectories ultimately converge on a common end-stage clinical phenotype characterized by widespread motor and systemic involvement. Regardless of the underlying explanation, these findings suggest that disease heterogeneity in ALS may be more effectively understood as a dynamic process rather than a collection of static subtypes. From both biological and clinical perspectives, modelling how patients transition from diverse early symptom profiles to increasingly overlapping late-stage presentations may provide a more informative framework for understanding disease progression than cross-sectional phenotyping alone.

### Clinical implications

#### Prhognosis

The identified clusters show distinct survival trajectories with potential clinical utility. The Sensory-Pain cluster had significantly longer survival than all other first-note clusters, while Motor-Bulbar, Motor-Tremor, and Motor-Respiratory clusters had similar, shorter survival profiles. In the all-notes analysis, Classic Motor patients had significantly shorter survival than both Autonomic-Respiratory and Nocturnal-Respiratory clusters. These survival differences, particularly those apparent from the first clinical note, could inform early care planning, prognostic discussions with patients and families, and the timing of interventions such as non-invasive ventilation and gastrostomy. Integrating NLP-derived symptom phenotypes with existing prognostic tools such as the ENCALS model and TRICALS risk score may further improve individualized outcome prediction.

#### Clinical trial stratification

Phenotype-based stratification could improve patient selection and enrichment for therapeutic trials, a recognized challenge in MND given the substantial heterogeneity in disease course [28]. The four first-note phenotypes may be particularly useful for early-stage trial enrichment, enabling recruitment of more homogeneous patient subgroups and potentially improving sensitivity to detect treatment effects. Data-driven phenotyping of this kind complements current eligibility criteria based on El Escorial or Gold Coast diagnostic categories, which were not designed to capture prognostic subgroups.

#### Healthcare resource planning

Beyond prognosis and trials, phenotype identification from routine EHR data could support multidisciplinary care planning. Patients in respiratory-predominant clusters may benefit from earlier referral to respiratory services, while bulbar-predominant patients may require prioritized speech and language therapy and nutritional support [29].

### Comparison with existing approaches

Previous data-driven clustering studies in MND have largely relied on structured registry data or small, carefully curated research datasets [14, 30, 11]. This approach differs in that it derives phenotypic features entirely from unstructured clinical text using NLP, making it scalable to any center with electronic health records and without requiring prospective data collection or bespoke research infrastructure. This is consistent with a growing body of work demonstrating the utility of NLP for phenotyping in other neurological conditions [31]. The use of UMLS-standardized concept extraction via MedCAT enables, in principle, cross-center replication using a common semantic framework. Unsupervised clustering of high-dimensional binary symptom data presents methodological challenges, and our use of latent class analysis is well-suited to this setting given the sparse, co-occurrence structure of the feature matrix.

### Strengths and limitations

This study has several notable strengths. The approach combines multi-method cohort confirmation, UMLS-based standardized concept extraction with negation and temporality detection, and sequential validation from first-note to all-notes analysis. The use of MedCAT, trained on large volumes of clinical text from comparable NHS settings, provides a robust and validated basis for concept extraction. The identified phenotypes mostly align with known MND pathophysiology and established clinical subtypes, supporting their biological plausibility. The real-world EHR setting also means that findings are grounded in routine clinical practice rather than highly selected research populations.

Several limitations should be acknowledged. This is a single-center study from a tertiary neurology referral center serving as a specialist hub for MND. While this provides access to a large, well-characterized cohort with detailed longitudinal clinical documentation, it may introduce selection bias towards more complex, atypical, or later-referred presentations; patients managed entirely in district general hospitals or primary care may exhibit different symptom profiles. Importantly, however, tertiary MND centers are precisely the settings in which NLP-based phenotyping pipelines of this kind are most deployable, as they accumulate the volume and depth of clinical documentation required for this approach. External validation in independent cohorts spanning both tertiary and community neurology settings, and ideally across different national healthcare systems is an aim of future work. The retrospective design is inherently subject to documentation biases: symptom recording reflects clinical priorities at the time of each encounter rather than systematic phenotypic assessment, and documentation practices may vary across clinicians and over time [32]. Ethnicity data were missing or recorded as unknown for 42.7% of the cohort. To mitigate the impact of this missingness, Cox proportional hazards models treated unknown ethnicity as an explicit category rather than excluding those patients, ensuring that survival estimates were not inadvertently biased by non-random missingness. The high rate of unknown ethnicity nonetheless limits the extent to which ethnicity-stratified analyses can be conducted, and future work should prospectively collect and harmonize ethnicity data using NHS Digital standards.

Despite filtering and negation detection, NLP extraction errors will persist to some degree. MedCAT has been extensively validated across multiple clinical domains and NHS datasets, with reported precision and recall exceeding 0.80 for a broad range of clinical entities [15], and its negation detection meta-annotations have been independently evaluated in clinical NLP benchmarks. The base model used here was further fine-tuned on our institutional clinical corpus using unsupervised training, improving adaptation to King’s College Hospital documentation conventions and terminology.

Binary symptom encoding, while necessary to construct a consistent patient-level feature matrix from heterogeneously documented clinical text, does not capture symptom severity, frequency, or temporal trajectory. In MND, the clinical significance of a symptom depends heavily on these dimensions: dyspnoea documented once in passing carries different prognostic weight than progressive dyspnoea documented across multiple encounters, yet both are represented identically in a binary matrix. This limitation is partly mitigated in the all-notes analysis, where symptoms that appear across multiple visits are more likely to be captured than those documented incidentally; however, severity gradients remain unrepresented. The choice of binary encoding was pragmatic: ordinal or continuous severity scores are rarely documented in a standardized format in routine free-text clinical notes, making their reliable extraction via NLP substantially more challenging. Future work should explore time-stamped, visit-level symptom matrices to capture longitudinal dynamics, and semi-structured clinical documentation, which would substantially improve the granularity available for NLP-based feature extraction.

Survival was calculated from the first recorded clinical encounter rather than symptom onset. This was a deliberate methodological choice as reliable symptom onset dates are not systematically documented in free-text clinical notes in our dataset and extracting them with sufficient accuracy and completeness via NLP was not feasible at scale. If clusters differ systematically in the delay between symptom onset and first clinic attendance, for example, if the Sensory-Pain cluster has a longer diagnostic journey, this could contribute to apparent survival differences. Structured data on genotype (including *C9orf72* and *SOD1* status), El Escorial or Gold Coast diagnostic category, and key clinical interventions (riluzole initiation, non-invasive ventilation, percutaneous endoscopic gastrostomy) were not systematically available for this cohort. These variables were not absent by design but reflect the realities of retrospective EHR data, as genetic testing was not universally performed or consistently recorded in structured fields during the study period, diagnostic categorization using formal criteria was not routinely coded in the electronic record, and intervention dates were documented inconsistently across clinical letter formats. Rather than conducting analyses on a substantially reduced subset with complete data, which would introduce further selection bias and reduce the generalizability of findings, we adopted a whole-cohort approach and acknowledge these as variables requiring prospective harmonization in future work.

### Future directions

Several avenues warrant further investigation. External validation of the identified phenotypes in independent MND cohorts represents an important next step. Longitudinal modelling of symptom trajectories, rather than cross-sectional symptom presence, may reveal additional subgroup structure and better capture disease progression dynamics. Integration of NLP-derived phenotypes with genetic, neuroimaging, and biomarker data (including neurofilament light chain levels) could provide a more complete characterization of MND subgroups and their biological underpinnings. Finally, a prospective evaluation of whether phenotype assignment at first presentation meaningfully influences clinical decision-making and patient outcomes would be required before clinical translation.

### Conclusions

We developed an NLP pipeline using MedCAT to discover symptom-based phenotypes in MND from EHR data. First-note analysis identified four initial presentations; all-notes analysis showed these reorganize into three longitudinal phenotypes with significantly different survival trajectories. These data-driven phenotypes extend beyond traditional onset-site classifications and may inform prognostication, early intervention, and clinical trial stratification.

## Data availability

Clinical data cannot be shared due to patient confidentiality.

## Code availability

Analysis code is available at https://github.com/yabdulle/mnd-symptom-phenotyping

## Acknowledgements

This work was supported by the UK Engineering and Physical Sciences Research Council (EP-SRC) [EP/Y035216/1] Centre for Doctoral Training in Data-Driven Health (DRIVE-Health) at King’s College London, with additional support from LifeArc. HW’s role in this research was partially funded by the Legal and General Group (research grant to establish the independent Advanced Care Research Centre at University of Edinburgh). The funders had no role in conduct of the study, interpretation, or the decision to submit for publication. The views expressed are those of the authors and not necessarily those of Legal & General. A.A.-C. is an NIHR Senior Investigator. A.A.-C. receives salary support from the National Institute for Health and Care Research (NIHR) Dementia Biomedical Research Unit at South London and Maudsley NHS Foundation Trust and King’s College London. A.I. is funded by South London and Maudsley NHS Foundation Trust, MND Scotland, Motor Neuron Disease Association, National Institute for Health and Care Research, Spastic Paraplegia Foundation, Rosetrees Trust, Darby Rimmer MND Foundation, the Medical Research Council (UKRI), Alzheimer’s Research UK and LifeArc. S.B is funded by the NHS England InnovateMD Fellowship.

## Author contributions

Y.A. performed the analyses, developed the pipeline, implemented the methods and drafted the manuscript. Y.A., A.I. and R.D. conceived the study. V.D. facilitated data access and curation. J.W., Y.K., S.B., Z.Y., and C.T. reviewed and provided critical feedback on the final manuscript. A.A-C., H.W., R.D., and A.I. supervised the study, provided guidance throughout, and contributed to manuscript revision. All authors approved the final version for submission.

