## Supplementary Material for "Symptom-based phenotype discovery in motor neuron disease using natural language processing of electronic health records" (from another version of this preprint: version 1, 2026-06-22)

Abdulle *et al (2026).*

**Contents**

1 Supplementary Methods	2
  1.1 NLP extraction parameters	2
  1.2 Clustering algorithm comparison	2
2 Supplementary Tables	3
  2.1 Extensive list of characteristing symptoms	3
      2.1.1 First note	3
      2.1.2 All notes	3
  2.2 Cox regression results by cluster	6
  2.3 Individual symptom Cox regression	6
3 Supplementary Figures	9
4 Data availability	12

### 1 Supplementary Methods

#### 1.1 NLP extraction parameters

MedCAT version 2.2.0 was used with a UMLS-based vocabulary. The following parameters were applied:

- Semantic type filter: T184 (Sign or Symptom)
- Confidence threshold: *≥* 0.5
- Negation detection: Enabled (MedCAT meta-annotation model)
- Context window: *±* 30 tokens
- Minimum patient prevalence: *≥* 5 patients
- Binary encoding: 1 (presence) or 0 (absence)

#### 1.2 Clustering algorithm comparison

We compared performance across multiple clustering algorithms on the all-notes symptom matrix (2,065 patients, 184 symptoms). All algorithms were evaluated at *k* = 3 (Table 1). Latent class analysis was selected as the primary method due to its probabilistic framework suited to binary data, interpretability, and strong performance. The low silhouette scores reflect high data sparsity (95.1% zeros), which is typical for symptom-based analyses in rare diseases.

Table 1: Clustering algorithm performance comparison (*k* = 3, all-notes matrix)

| **Algorithm** | **Silhouette** | **Calinski-Harabasz** | **BIC** |
| --- | --- | --- | --- |
| Latent Class | 0.16 | 188.9 | 86,045 |
| Bernoulli Mixture | 0.16 | 187.9 | – |
| K-means (raw) | 0.13 | 202.6 | – |
| K-means (NMF) | 0.09 | 164.4 | – |
| NMF-Cosine | 0.07 | 150.3 | – |
| Spectral | 0.06 | 162.8 | – |
| Agglomerative | *−*0.01 | 1.4 | – |

### 2 Supplementary Tables

#### 2.1 Extensive list of characterizing symptoms

##### 2.1.1 First note

**Table 2: Symptoms in the first-note binary matrix after filtering (n = 76 symptoms, 2,065 patients).**

| **Symptoms** | |
| --- | --- |
| Aching Muscles | Action Tremor |
| Apnoeic | Articular Pain |
| Ataxia of Gait | Bladder Symptoms |
| Charming | Cramp and Spasm |
| Decreased Food Appetite | Difficulty Opening Bowels |
| Difficulty Sleeping | Dysdiadochokinesia |
| Dysphagia | Equilibration Disorder |
| Eruption of Skin | Excessive Daytime Sleepiness |
| Fasciculations | Fasciculations Benign |
| Fatigue | Fatigueability |
| Floppy | General Illness |
| Generalized Muscle Weakness | Giving Way |
| Headache | Heart Problems |
| Hoarseness Throat Symptom | Increased Respiratory Depth |
| Ischaemic Heart Disease Angina | Lethargic |
| Lightning Pains | Malaise |
| Motor Neuron Signs | Muscle Cramps in Lower Leg |
| Muscle Weakness | Neurological Symptom Findings |
| Nonspecific Pain in the Lumbar Region | Nonspecific Pain in the Neck Region |
| Ns Neck Stiffness | Numbness |
| Of Diarrhoea | Of Nausea |
| Of Sneezing | Of Snoring |
| Out of Breath | Pain |
| Pain in Back | Paraesthesia |
| Passage of Watery Stools | Precipitated By Exercise |
| Presenting Complaint | Progressive Limb Weakness |
| Progressive Spastic Paraparesis | Pruritic Dermatitis |
| Quadriparesis | Radiating Pains |
| Respiratory Symptoms | Resting Tremor |
| Rigidity Muscular | Sensory Changes |
| Sensory Discomfort | Sensory Loss |
| Shortness of Breath | Signs and Symptoms |
| Slow Physical Movement | Soboe Shortness of Breath on Exertion |
| Spastic Paraparesis | Spasticity |
| Stiffness | Tendency to Fall |
| Tremor | Unspecified Visual Loss |
| Upper Motor Neuron Signs | Weak Cough |
| Weakness | Wheezing |

##### 2.1.2 All notes

**Table 3: Symptoms in the all-notes binary matrix after filtering (n = 183 symptoms, 2,065 patients).**

| **Symptom** | **Symptom** |
| --- | --- |
| Abdominal Bloating | Abdominal Crampy Pains |
| Aching Muscles | Aching Sensation Quality |
| Action Tremor | Agitated Behaviour |
| Aphasic | Apnoeic |
| Apraxic | Arthralgia of Ankle |
| Articular Pain | Ataxia of Gait |
| Back Severe Pain | Blackout Symptom |
| Bladder Symptoms | Blushing Function |
| Breathing Symptoms | Burning Chest Pains |
| Change in Bowel Pattern | Charming |
| Chest Symptoms Complaints | Choking on Liquid |
| Chronic Back Pain | Chronic Cough |
| Cogwheel Muscle Rigidity | Coughing Up Blood |
| Coughing Up Phlegm | Cramp and Spasm |
| Cramp in Lower Leg Associated with Rest | Decreased Food Appetite |
| Decreased Grip Strength | Despondent |
| Diarrhoea | Difficulties with Micturition |
| Difficulty Opening Bowels | Difficulty Sleeping |
| Difficulty Swallowing Getting Worse | Distressing Symptoms |
| Disturbance of Orientation | Dribbling Saliva |
| Dysdiadochokinesia | Dysphagia |
| Early Satiety | Edema of Foot Finding |
| Enlarged Lymph Nodes Unspecified | Equilibration Disorder |
| Eructation | Eruption of Skin |
| Excessive Daytime Sleepiness | Extremity Stiffness |
| Eyes Weakness | Fasciculations |
| Fasciculations Benign | Fatigue |
| Fatigueability | Feeling Abnormal |
| Feeling Emotional Such As Angry Or Upset | Feeling of Chest Tightness |
| Floppy | Gasping Respiration |
| Gastrointestinal Symptoms | General Body Deterioration |
| General Feeling of Illness | General Illness |
| General Symptoms | Generalized Muscle Weakness |
| Giddy Mood | Giving Way |
| Grimacing | Hand Symptoms |
| Headache | Headache on Entire Left Side |
| Headaches Severe | Heart Problems |
| Hesitancy of Micturition | Hoarseness Throat Symptom |
| Increased Respiratory Depth | Increased Shortness of Breath |
| Inguinal Pain | Intermittent Pain |
| Involuntary Quiver | Irritation Nerve Root |
| Ischaemic Heart Disease Angina | Jaundiced |
| Jaw Stiffness | Lack of Memory |
| Lead Pipe Muscle Rigidity | Leg Spasm |
| Legs Feel Heavy Symptom | Lethargic |
| Light Headedness | Lightning Pains |
| Malaise | Metallic Taste in the Mouth |
| Motor Neuron Signs | Muscle Cramps in Lower Leg |
| Muscle Weakness | Nausea |
| Neck | Neck Discomfort |
| Neurobehavioral Manifestations | Neurological Symptom Findings |
| Neuromuscular Weakness | No Adverse Effect |
| No New Symptoms | Non Productive Cough |
| Nonspecific Pain in the Lumbar Region | Nonspecific Pain in the Neck Region |
| Ns Neck Stiffness | Numbness |
| Numbness and Tingling | Oedema Pitting |
| Out of Breath | Pain |
| Pain in Back | Pain in Bilateral Legs |
| Pain in Calf | Pain in Left Upper Limb |
| Pain in the Lumbar Spine | Pain of Right Knee Region |
| Pain on Swallowing | Painful Hips |
| Pains Nerve | Paraesthesia |
| Passage of Watery Stools | Pharyngeal Dryness |
| Pleuritic Chest Pain | Polymyalgia |
| Precipitated By Exercise | Previously Well |
| Problem with Kidney | Progressive Limb Weakness |
| Progressive Spastic Paraparesis | Pruritic Dermatitis |
| Pyrexial | Quadriparesis |
| Radiating Pains | Reduced Reflexes |
| Respiratory Symptoms | Resting Tremor |
| Rigidity Muscular | Rigidity of Limbs |
| Romberg Sign Finding | Sensation of Suffocation |
| Sensory Changes | Sensory Discomfort |
| Sensory Loss | Shaking of Hands |
| Shallow Respirations | Sharp Pain Chest |
| Shortness of Breath | Shoulder Region Pain |
| Signs and Symptoms | Sleep Symptoms |
| Slow Physical Movement | Sneezing |
| Snoring | Soboe Shortness of Breath on Exertion |
| Spastic Paraparesis | Spasticity |
| Stiffness | Stomach Pain Ache |
| Symptoms of Depression | Systemic Symptoms |
| Tendency to Fall | Tremor |
| Tremors Coarse | Tremors Fine |
| Tremors Intermittent | Tremors Pill Rolling |
| Tremors in Hands | Unspecified Skin Changes |
| Unspecified Visual Loss | Upper Motor Neuron Signs |
| Weak Cough | Weak Grip |
| Weak Respiration | Weakness |
| Weakness Shoulder Right Only | Weakness Worsened |
| Weakness in Fingers | Weakness of Bilateral Upper Extremities |
| Wheezing |  |

#### 2.2 Cox regression results by cluster

Table 4: Cluster-level hazard ratios (first-note analysis)

| **Cluster** | **Phenotype** | **HR** | **95% CI** | **p-value** | **Sig.** |
| --- | --- | --- | --- | --- | --- |
| 0 | Motor-Bulbar | 1.54 | 1.24–1.92 | *<*0.001 | Yes |
| 1 | Motor-Tremor | 1.32 | 1.01–1.73 | 0.042 | Yes |
| 2 | Sensory-Pain | 1.00 | ref | – | – |
| 3 | Motor-Respiratory | 1.31 | 0.97–1.78 | 0.080 | No |

In the first-note analysis (n = 816), cluster assignment was significantly associated with survival (adjusted Cox likelihood-ratio test χ²(3) = 15.7, p = 0.001). Kaplan–Meier curves also differed significantly across clusters (log-rank p = 0.0001).

Table 5: Cluster-level hazard ratios (all-notes analysis)

| **Cluster** | **Phenotype HR** | **95% CI** | **p-value** | **Sig.** |
| --- | --- | --- | --- | --- |
| 0 | Autonomic-Respiratory 0.99 | 0.83–1.19 | 0.950 | No |
| 1 | Nocturnal-Respiratory 1.00 | ref | – | – |
| 2 | Classic Motor 1.31 | 1.12–1.54 | 0.001 | Yes |

In the all-notes analysis (n = 1,998), cluster assignment remained significantly associated with survival after adjustment (Cox LRT χ²(2) = 24.6, p < 0.001; log-rank p < 0.001).

#### 2.3 Individual symptom Cox regression

The following cox models assess the association between survival and the five most frequent symptoms within each phenotype cluster.

Table 6: Prevalent symptom hazard ratios, First-Note - Motor-Bulbar (n=355)

| **Symptom** | **HR** | **95% CI** | **p-value** | **Sig.** | **% Patients** |
| --- | --- | --- | --- | --- | --- |
| Weakness | 0.93 | 0.63–1.39 | 0.740 | No | 91.5 |
| Fasciculations | 1.38 | 0.90–2.12 | 0.141 | No | 89.3 |
| Spasticity | 0.96 | 0.74–1.26 | 0.787 | No | 29.0 |
| Shortness of Breath | 1.16 | 0.88–1.53 | 0.285 | No | 26.5 |
| Dysphagia | 0.94 | 0.70–1.27 | 0.689 | No | 20.6 |

Table 7: Prevalent symptom hazard ratios, First-Note - Motor-Tremor (n=140)

| **Symptom** | **HR** | **95% CI** | **p-value** | **Sig.** | **% Patients** |
| --- | --- | --- | --- | --- | --- |
| Fasciculations | NE*^†^* | — | — | — | 100.0 |
| Weakness | 0.97 | 0.61–1.53 | 0.899 | No | 73.6 |
| Tremor | 0.64 | 0.43–0.98 | 0.039 | Yes | 46.4 |
| Spasticity | 1.04 | 0.67–1.59 | 0.873 | No | 34.3 |
| Slow Physical Movement | 1.29 | 0.72–2.32 | 0.391 | No | 16.4 |

*^†^* Hazard ratio not estimable for symptoms present in *≥*98% of patients within the cluster (insufficient variation for Cox regression).

Table 8: Prevalent symptom hazard ratios, First-Note - Sensory-Pain (n=198)

| **Symptom** | **HR** | **95% CI** | **p-value** | **Sig.** | **% Patients** |
| --- | --- | --- | --- | --- | --- |
| Weakness | 1.25 | 0.75–2.07 | 0.393 | No | 80.8 |
| Fasciculations | 1.73 | 1.01–2.97 | 0.046 | Yes | 79.8 |
| Pain | 1.16 | 0.79–1.70 | 0.455 | No | 51.0 |
| Sensory Loss | 0.71 | 0.48–1.05 | 0.086 | No | 40.9 |
| Paraesthesia | 1.18 | 0.73–1.94 | 0.498 | No | 16.2 |

Table 9: Prevalent symptom hazard ratios, First-Note - Motor-Respiratory (n=123)

| **Symptom** | **HR** | **95% CI** | **p-value** | **Sig.** | **% Patients** |
| --- | --- | --- | --- | --- | --- |
| Shortness of Breath | NE*^†^* | — | — | — | 100.0 |
| Excessive Daytime Sleepiness | NE*^†^* | — | — | — | 99.2 |
| Decreased Food Appetite | 0.35 | 0.11–1.07 | 0.065 | No | 96.7 |
| Presenting Complaint | 0.78 | 0.27–2.30 | 0.657 | No | 95.9 |
| Spasticity | 0.62 | 0.23–1.67 | 0.345 | No | 94.3 |

*^†^* Hazard ratio not estimable for symptoms present in *≥*98% of patients within the cluster (insufficient variation for Cox regression).

Table 10: Prevalent symptom hazard ratios, All-Notes Autonomic- Respiratory (n=472)

| **Symptom** | **HR** | **95% CI** | **p-value** | **Sig.** | **% Patients** |
| --- | --- | --- | --- | --- | --- |
| Weakness | 1.03 | 0.48–2.22 | 0.943 | No | 97.9 |
| Fasciculations | 1.27 | 0.75–2.14 | 0.378 | No | 94.7 |
| Shortness of Breath | 1.25 | 0.90–1.75 | 0.183 | No | 84.7 |
| Headache | 0.81 | 0.62–1.06 | 0.125 | No | 81.8 |
| Pain | 0.64 | 0.50–0.81 | *<*0.001 | Yes | 72.7 |

Table 11: Prevalent symptom hazard ratios, All-Notes Nocturnal-Respiratory (n=338)

| **Symptom** | **HR** | **95% CI** | **p-value** | **Sig.** | **% Patients** |
| --- | --- | --- | --- | --- | --- |
| Excessive Daytime Sleepiness | NE*^†^* | — | — | — | 100.0 |
| Shortness of Breath | NE*^†^* | — | — | — | 100.0 |
| Headache | 0.75 | 0.24–2.39 | 0.627 | No | 98.2 |
| Decreased Food Appetite | NE*^†^* | — | — | — | 98.2 |
| Weak Cough | 1.16 | 0.57–2.37 | 0.684 | No | 95.6 |

*^†^* Hazard ratio not estimable for symptoms present in *≥*98% of patients within the cluster (insufficient variation for Cox regression).

Table 12: Prevalent symptom hazard ratios, All-Notes Classic Motor (n=1188)

| **Symptom** | **HR** | **95% CI** | **p-value** | **Sig.** | **% Patients** |
| --- | --- | --- | --- | --- | --- |
| Fasciculations | 1.26 | 0.98–1.62 | 0.067 | No | 90.2 |
| Weakness | 1.01 | 0.81–1.26 | 0.909 | No | 89.7 |
| Pain | 0.74 | 0.64–0.86 | *<*0.001 | Yes | 33.8 |
| Spasticity | 0.91 | 0.78–1.06 | 0.212 | No | 30.0 |
| Shortness of Breath | 1.53 | 1.32–1.78 | *<*0.001 | Yes | 25.3 |

### 3 Supplementary Figures

Figure 1: UMAP visualization of first-note symptom profiles. Points represent individual patients colored by cluster assignment.

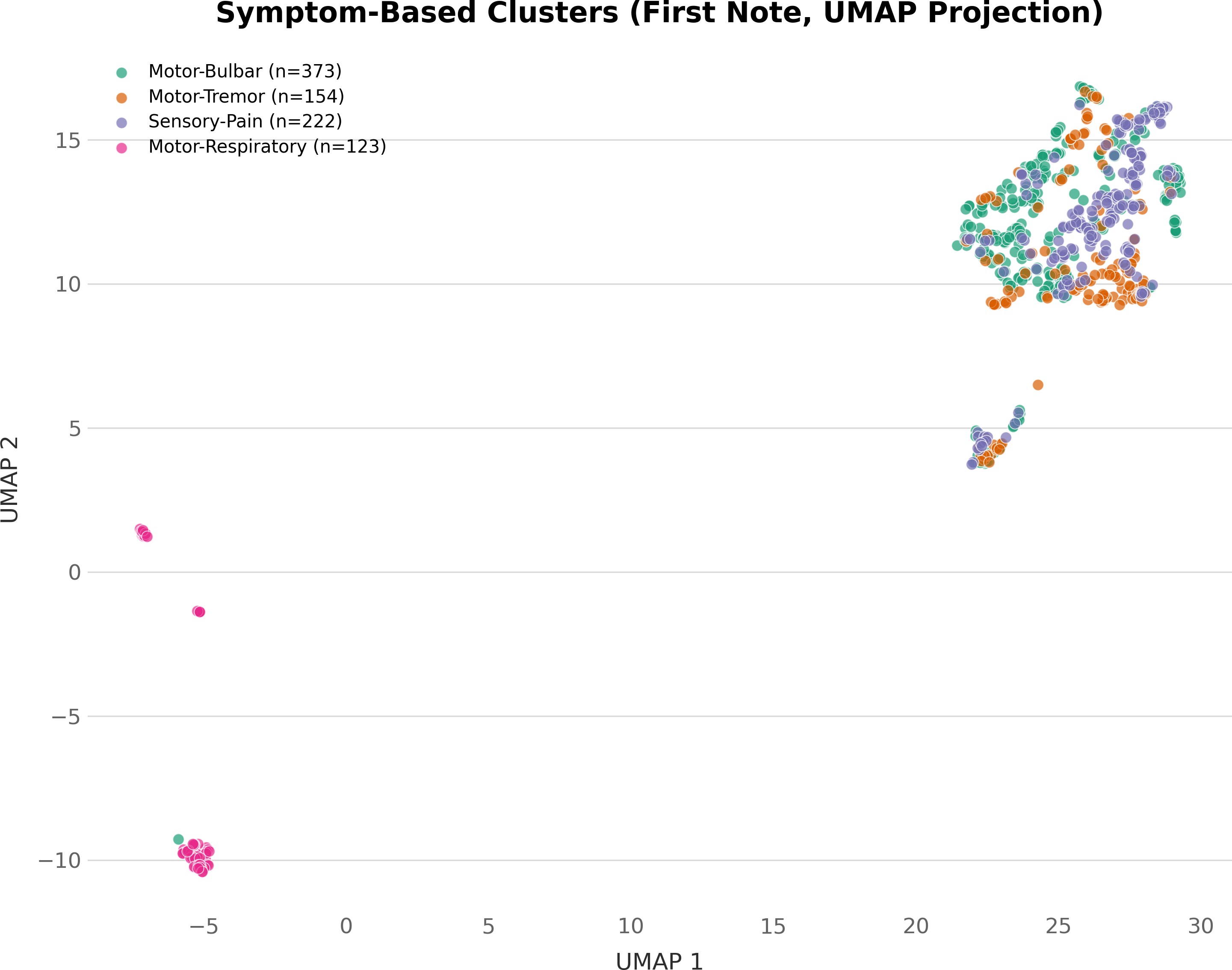

Figure 2: UMAP visualization of all-notes symptom profiles.

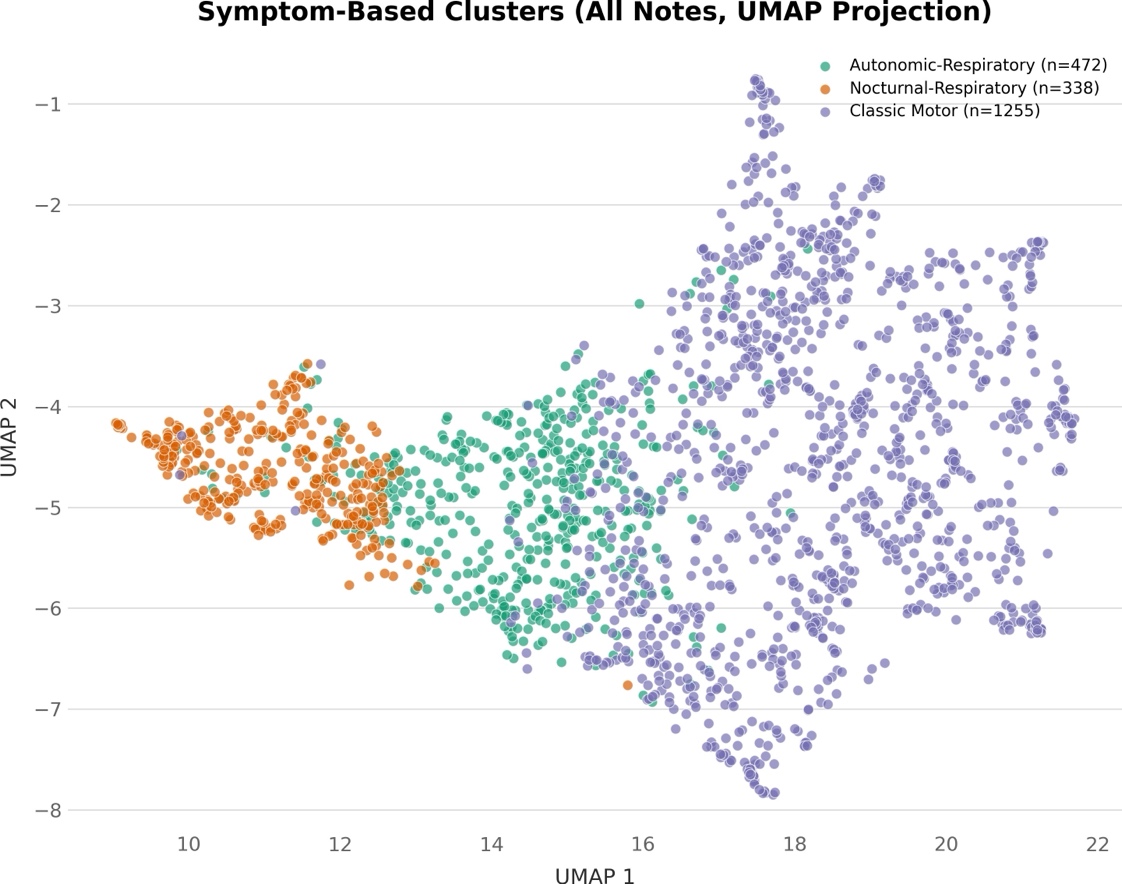

Figure 3: Clustering method comparison for first-note analysis.

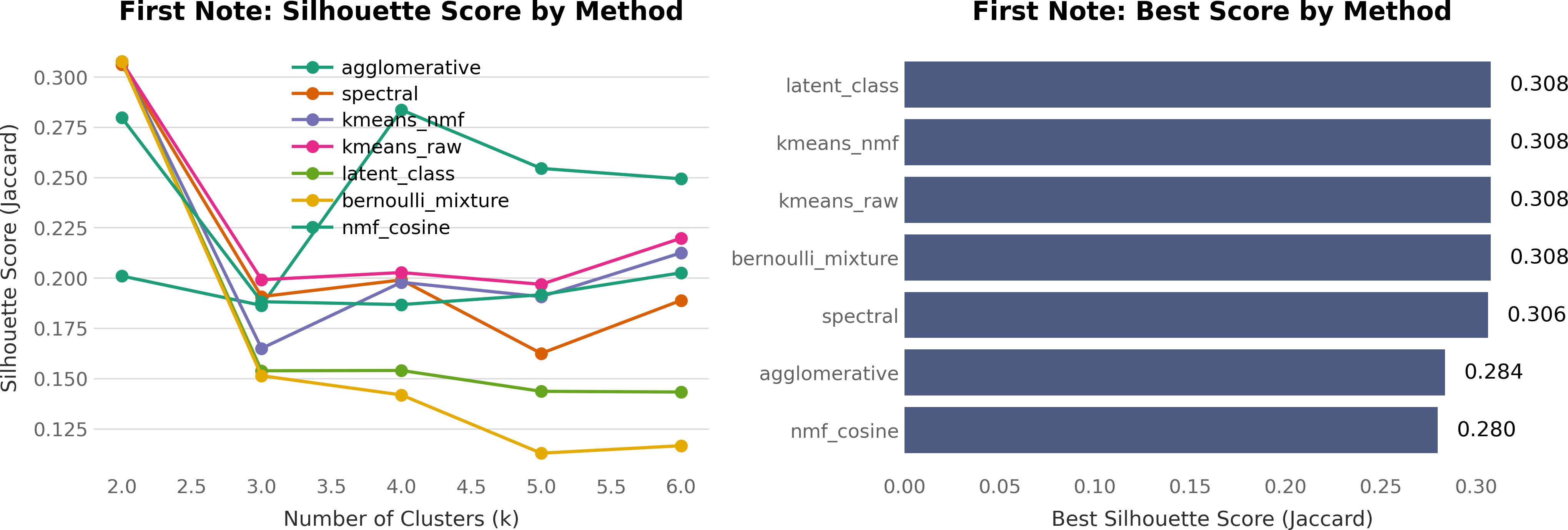

Figure 4: Clustering method comparison for all-notes analysi
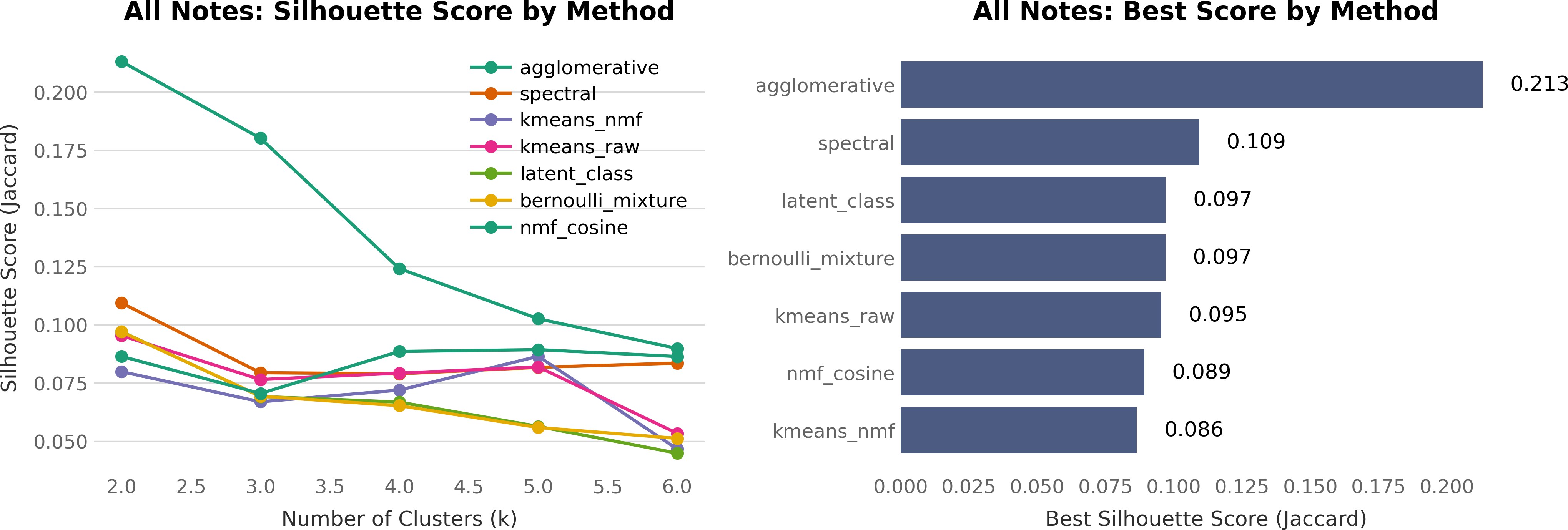

### 4 Data availability

Clinical data cannot be shared due to patient confidentiality and data governance requirements. Analysis code is available at <https://github.com/yabdulle/mnd-symptom-phenotyping>.
